# THE INTESTINAL AND ORAL MICROBIOMES ARE ROBUST PREDICTORS OF COVID-19 SEVERITY THE MAIN PREDICTOR OF COVID-19-RELATED FATALITY

**DOI:** 10.1101/2021.01.05.20249061

**Authors:** Doyle V. Ward, Shakti Bhattarai, Mayra Rojas-Correa, Ayan Purkayastha, Devon Holler, Ming Da Qu, William G. Mitchell, Jason Yang, Samuel Fountain, Abigail Zeamer, Catherine Forconi, Gavin Fujimori, Boaz Odwar, Caitlin Cawley, Beth A. McCormick, Ann Moormann, Mireya Wessolossky, Vanni Bucci, Ana Maldonado-Contreras

## Abstract

The reason for the striking differences in clinical outcomes of SARS-CoV-2 infected patients is still poorly understood. While most recover, a subset of people become critically ill and succumb to the disease. Thus, identification of biomarkers that can predict the clinical outcomes of COVID-19 disease is key to help prioritize patients needing urgent treatment. Given that an unbalanced gut microbiome is a reflection of poor health, we aim to identify *indicator species* that could predict COVID-19 disease clinical outcomes. Here, for the first time and with the largest COVID-19 patient cohort reported for microbiome studies, we demonstrated that the intestinal and oral microbiome make-up predicts respectively with 92% and 84% accuracy (Area Under the Curve or AUC) severe COVID-19 respiratory symptoms that lead to death. The accuracy of the microbiome prediction of COVID-19 severity was found to be far superior to that from training similar models using information from comorbidities often adopted to triage patients in the clinic (77% AUC). Additionally, by combining symptoms, comorbidities, and the intestinal microbiota the model reached the highest AUC at 96%. Remarkably the model training on the stool microbiome found enrichment of *Enterococcus faecalis*, a known pathobiont, as the top predictor of COVID-19 disease severity. *Enterococcus faecalis* is already easily cultivable in clinical laboratories, as such we urge the medical community to include this bacterium as a robust predictor of COVID-19 severity when assessing risk stratification of patients in the clinic.

## INTRODUCTION

An estimated 20% of individuals infected with SARS-CoV-2 require hospitalization, with a subset of patients requiring intensive care. Why some individuals become deathly ill while others don’t is still unknown. Despite the rollout of vaccination campaigns against SARS-CoV-2, the threat of this infection is ongoing. As hospitals worldwide are challenged with episodic resurgences of patients with COVID-19 disease, there is an urgent need for pragmatic yet accurate risk stratification biomarkers that could predict which patients are high risk for progression to severe disease and death. Precise risk stratification protocols can help justify resource allocation, if faced with limitations, and guide staffing decisions for efficient patient management.

In an effort to develop such a protocol, the International Severe Acute Respiratory and emerging Infections Consortium (ISARIC) World Health Organization (WHO) described in September 2020, a comprehensive risk stratification tool for SARS-CoV-2 hospitalized patients: 4C Mortality score ^1^. The 4C Mortality Score included eight variables: age, sex, number of comorbidities, respiratory rate, peripheral oxygen saturation, level of consciousness, urea level, and C reactive protein. However, this scoring system has only 79% of accuracy ^1^. That is, out of 10 COVID-19 patients, the 4C Mortality Score will fail to identify 3 patients with high risk of fatality. Thus, a more precise method is critically needed to forecast hospital capacity during this pandemic. It has been established that patients with SARS-Cov-2 infection exhibit gut microbiome dysbiosis when compared to healthy individuals (reviewed ^2^). More recently, a total of 23 bacterial taxa was found to be strongly associated with disease severity among hospitalized COVID-19 patients ^3^.

Here, we capitalized on a robust and validated predictive analytic and computational framework developed by us ^4-6^ to define and model complex interactions between the microbiota, clinical variables, and disease severity. Hence, we discovered oral and intestinal bacteria species that can be used to accurately predict fatality of COVID-19 hospitalized patients.

## RESULTS

### Fatality of patients with SARS-CoV-2 infection is predicted by respiratory severe symptoms

We enrolled 69 SARS-CoV-2 PCR positive patients with moderate or severe symptoms. In accordance with the categorization of disease severity used in the hospital, those requiring more than 4 L of oxygen (at the time of sample collection) were considered patients with severe symptoms; conversely, patients needing less than 4L of oxygen were categorized as having moderate symptoms. Out of 69, we included in the analysis 63 participants with complete medical records including disease outcomes (Table 1). As seen in table 1, there were no differences between the two groups in age, body mass index (BMI), sex, race, smoking status, or antibiotic administration during hospitalization. However, we observed significant differences in the duration of the hospital stay: patients with severe symptoms had an average of ∼6 more days in the hospital than patients exhibiting moderate symptoms. Additional information about COVID-19 symptoms and comorbidities included in the subsequent analyses are detailed in Supplementary Table 1.

**Table 1.** Characteristics of COVID-19 hospitalized patients recruited in the study from April to June 2020.

|  | Moderate (n=32) | Severe (n=31) | *p-value* |
| --- | --- | --- | --- |
| *Demographics* |  |  |  |
| Age (years) | 70.53 ± 15.86 | 70.58 ± 14.08 | 0.8* |
| BMI | 29.22 ± 9.19 | 30.58 ± 8.82 | 0.2* |
| Female (%) | 16 (50%) | 10 (30%) | 0.2<sup>†</sup> |
| *Race* |  |  |  |
| White (%) | 24 (75%) | 19 (59.37%) | 0.2<sup>^</sup> |
| Black or African American (%) | 4 (12.50%) | 3 (9.37%) |  |
| Hispanic or Latino (%) | 4 (12.50%) | 8 (25%) |  |
| Asian (%) | 0 (0.00%) | 2 (6.25%) |  |
| *Smoking status* |  |  |  |
| Current | 2 (6.25%) | 4 (12.50%) | 0.6<sup>^</sup> |
| Former | 11 (34.38%) | 11 (34.37%) |  |
| Never | 19 (59.38%) | 16 (50%) |  |
| Antibiotics treatment | 26 (81.25%) | 29 (90.62%) | 0.2<sup>^</sup> |
| Days in the hospital | 15.5 + 10.39 | 21.27 + 12.73 | **0.045\*** |

\*Mann Whitney, unpaired t-test
<sup>†</sup>Fisher T-test
<sup>^</sup> Chi-square

We applied Random Forest Classification to determine which of the 68 clinical covariates (Table 1 and Supplementary Table 1) plus severity of symptoms as described above could predict COVID-19 fatality on the patient cohort recruited for this study. We used the Boruta algorithm to perform feature selection and identify all the relevant clinical covariates. We found that a combination of clinical covariates which includes disease severity was able to predict a patient succumbing to COVID-19 disease with an 89% accuracy (Area Under the Curve-Receiving Operating Curve, AUC-ROC) on leave-one-out cross validation data (Figure 1). In fact, the modeling identified the hospital classification of disease severity at the 4L of oxygen requirement cut-off as the main factor predicting a patient’s fatality based on the Random Forest Classification estimated Variable Importance values ^7^ with AUC-ROC dropping to 84% when the disease severity variable was omitted (Figure 1A and B). In addition, we found that other clinical variables were also more common in patient that had poor outcomes (Figure 1C and D).

**Figure 1.**
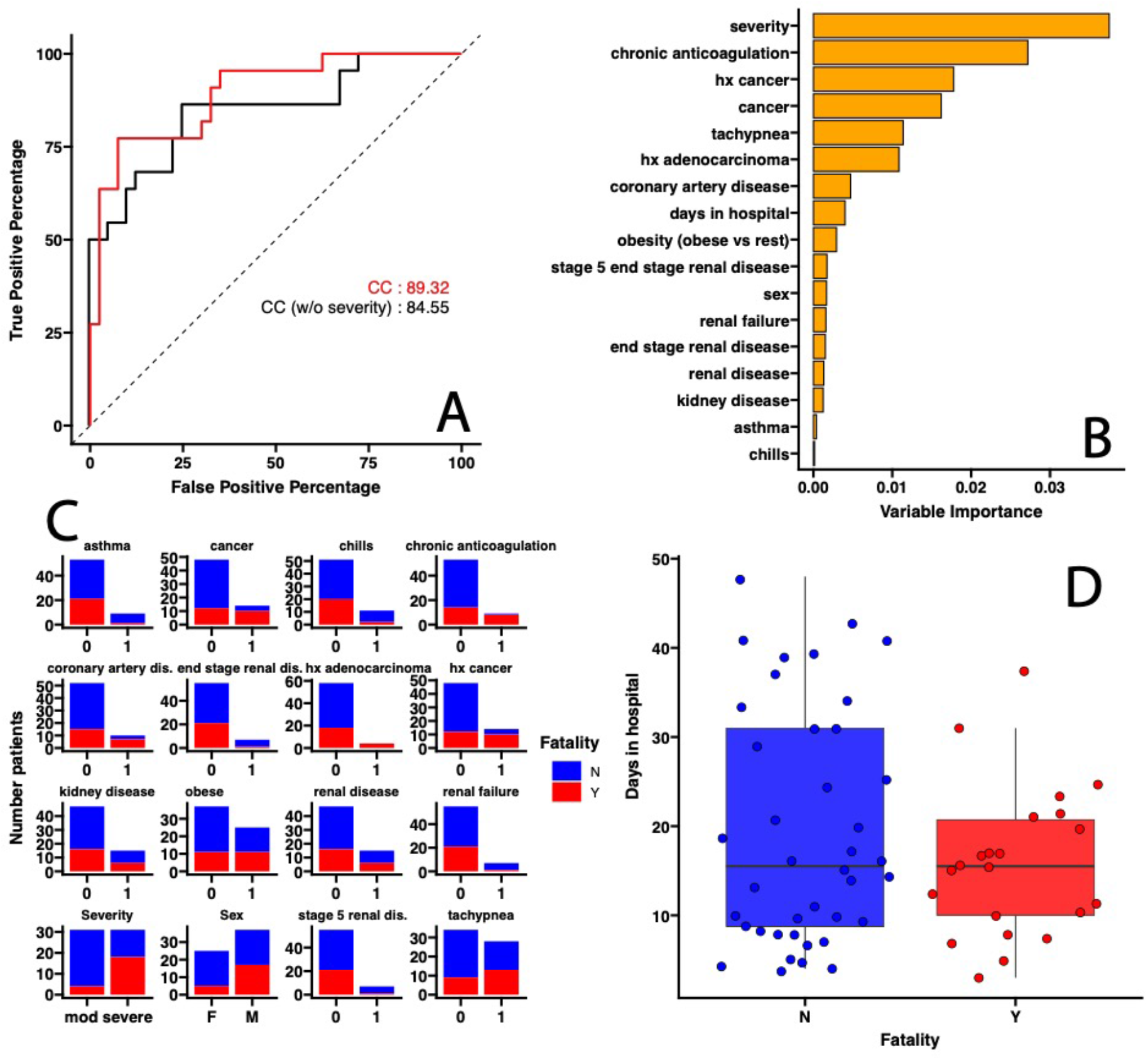
COVID-19 fatality is predicted by severity of respiratory symptoms and other comorbidities commonly used to triage patients. (A) Area Under the Curve-Receiving Operating Curve (AUC-ROC) for leave-one-out cross-validation evaluating prediction of accuracy of COVID-19 fatality. Red lines correspond to the model including all the clinical covariates (CC), black line correspond to the model including all the clinical covariates except disease severity (CC, no Severity). (B) Covariates selected by the Random Forest Classification model ranked according to their importance in classifying fatality as a disease outcome. (C) For categorical covariates (Yes=1, No=0) the number of patients out of the 63 included in the analyses within a specific category were colored by outcome (Survived, in blue; Died, in red). (D) For numerical variable, whisker plots (median, box interquartile range, 5^th^ and 9^th^ percentile for lines) are used with each solid dot corresponding to a single patient. (BH adjusted *p value* < 0.05)

Together, these suggest that prediction of COVID-19 outcomes is improved when taking into account respiratory symptoms, namely requiring more than 4 L of oxygen, along with other clinical variables commonly used to do so.

### Disease severity is accurately predicted by stool or oral microbiome

Studies have shown that viral lung infections, including SARS-CoV-2, have a lasting effect on the gut microbiota ^3,8-11^. Therefore, the composition of the microbiota may contain information that is not directly measured or quantified by clinicians when triaging COVID-19 patients. We therefore decided to compare the ability in predicting COVID-19 disease severity (the main predictor of fatality) by the gastrointestinal and oral microbiome compared to commonly measured clinical covariates. We applied Random Forest Classification to predict severe *vs*. moderate symptoms as a function of (i) only clinical variables (Table 1 and Supplementary Table 1), (ii) intestinal microbiome composition, (iii) oral microbiome composition, (iv) clinical variables and intestinal microbiome composition combined, and (v) clinical variables and oral microbiome composition combined. In the clinical variables we included age and BMI as additional explanatory variables to control for their effect. Here, we included a total of 62 patients that provided either or both stool and tongue samples. The model trained only on clinical variables was found to predict COVID-19 disease severity with 75.55% AUC-ROC using leave-one-out cross-validation (Figure 2A) comparable to what have been previously reported ^1^. This model determined as significant predictors of COVID-19 disease severity, 10 clinical variables: hypercholesteremia, race (Latino), coronary artery disease, asthma, obesity, hypoxic respiratory stress, tachypnea, days in hospital, thrombosis, and sex (male).

**Figure 2.**
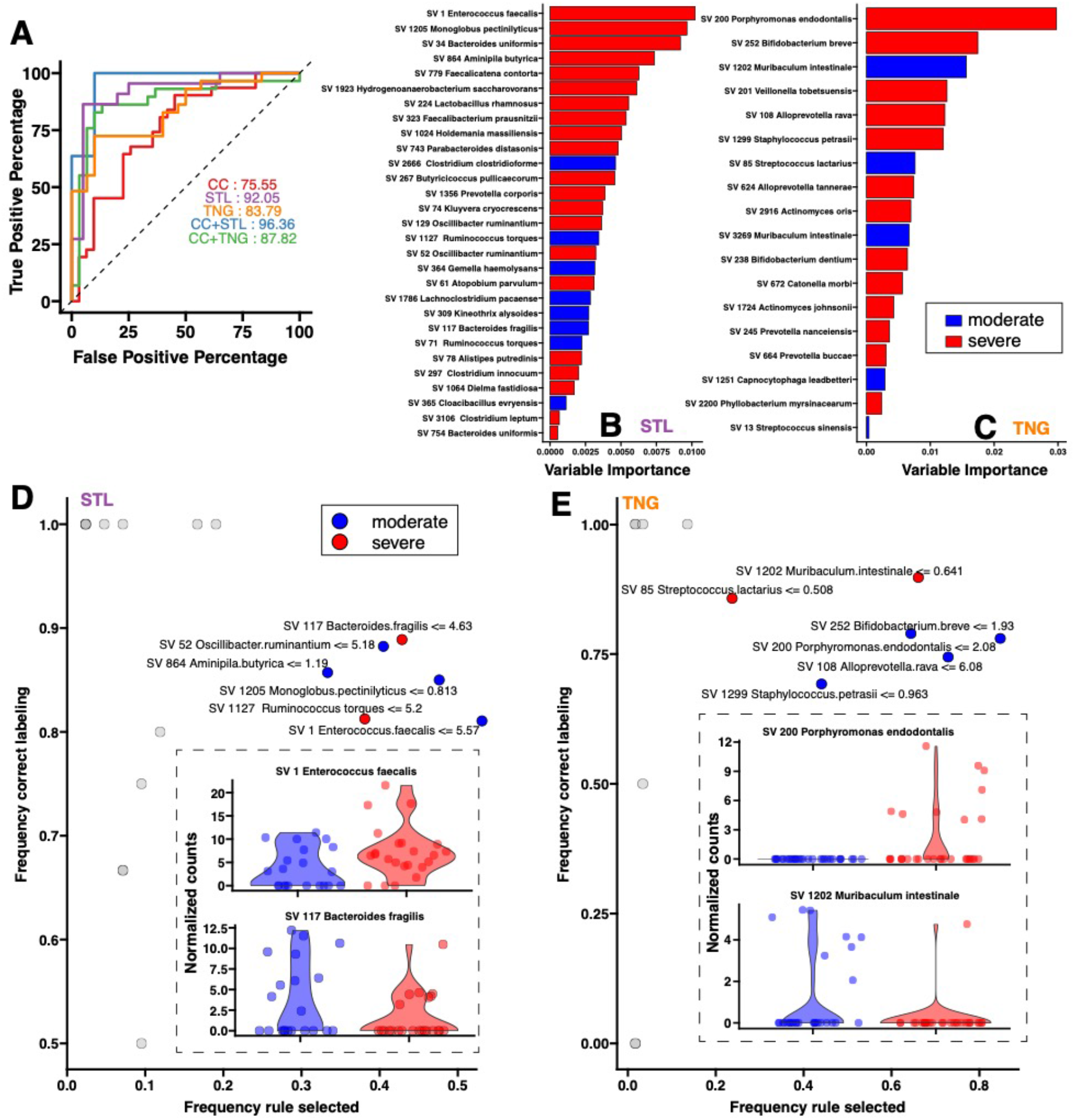
Stool and oral microbiotas predict COVID-19 disease severity with significant greater accuracy compared to clinical variables alone. (A) Receiving Operating Curves (ROCs) for leave-one-out cross-validation evaluating prediction of accuracy of severity for different models. The models accounting for stool microbiome (STL) or oral microbiome (TNG) predict severity with greater accuracy than models trained solely with clinical covariates (CC). Addition of clinical covariates to microbiome variables increase predictive ability to 96.36% AUC. Bacterial species from the microbiome selected by the Random Forest models for STL (B) and TNG (C) ranked according to their importance in classifying COVID-19 disease severity. We ran Local Interpretable Model-agnostic Explanation (LIME) analysis to determine direction of the COVID-19 – microbiota associations. LIME identifies logical rules that best separates between the two outcome groups (moderate = blue, severe = red). For each rule, the frequency of each rule being selected *vs*. the frequency of that rule predicting the true category across all the cross-validation runs are displayed. Rules on *Enterococcus faecalis* in the STL set (D) and on *Porphyromonas endodontalis* in the TNG (E) set are found to have primary discriminatory power in classifying COVID-19 disease severity. While the model points to enrichment of these two pathobionts respectively in the intestinal and in the oral mucosa as major predictors of disease severity; conversely, reduction of *Bacteroides fragilis* and *Moribaculum intestinale* are also selected as primary COVID-19 disease severity indicators.

We then performed the same computational analysis but this time assessing the intestinal microbiota (measured in stool, referred as STL) or the oral microbiota (measured in tongue, referred as TNG) as predictors. These models were able to predict COVID-19 disease severity with 92.05% and 83.79% AUC-ROC, respectively. These represent an improvement in prediction accuracy of 122% for intestinal, and 111% for oral microbiota compared to what is achieved using clinical variables alone (Figure 2A). Furthermore, combining clinical variables and STL or TNG microbiomes abundances improve prediction ability, with the highest AUC-ROC of 96.36%. This analysis suggests that the intestinal and oral microbiotas individually can provide a more accurate and robust biomarker of disease severity that may not be quantifiable by other clinical variables assessed during patient triage. Additional metrics on prediction ability of the different models are reported in the Supplementary Table 2.

### Abundance of *indicator species* is a predictor of COVID-19 disease severity

Given the time, expertise, and resources necessary for the analysis and interpretation of microbiota data, we further investigated indicator species of the oral and intestinal microbiota that can be easily cultured in clinical laboratory settings ^12,13^ and can be added as a test for risk stratification of COVID-19 patients. In ecological research, indicator species have been defined as those that serve as a surrogate measures of the health or lack thereof of an entire ecosystem^14,15^. We reasoned that indicator species within the microbiota ecosystem could act as surrogate markers of COVID-19 disease severity. Again, after performing feature selection with Boruta, we ran Random Forest Classification using only the Boruta-selected features for every model in Figure 2A. After features ranking based on applied Random Forest Classification-estimated Variable Importance values, we found that the top three bacterial species with the strongest likelihood to predict COVID-19 disease severity from the intestinal microbiome were *Bacteroides uniformis (Bacteroides/ Bacteroidia), Enterococcus faecalis (Firmicutes/ Bacilli)*, and *Monoglobus pectinilyticus (Firmicutes/ Clostridia)*; and from the oral microbiome were *Porphyromonas endodontalis (Bacteroides/ Bacteroidia), Veillonella tobetsuensis (Firmicutes/ Negavicutes)*, and *Bifidobacterium breve* (Actinobacteria/ Actinobacteria. Figure 2B and C).

To determine the direction (positive/negative) of the identified microbial abundances related to COVID-19 severity we then ran Local Interpretable Model-agnostic Explanation (LIME) analysis ^16^. LIME trains a local surrogate model that can be used to explain the predictions of a ‘black-box’ machine learning model such as Random Forest thus providing confidence that the model will perform well on real-world data, crucial for medical decision making. In our context LIME identifies human-interpretable rules on the microbiome that discriminate between patients with moderate or severe COVID-19 symptoms. LIME analyses predicted that reduced abundance in moderately ill patients (and *vice versa* enrichment in severely ill patients) of the known gastrointestinal pathobiont *Enterococcus faecalis* and of the oral pathobiont *Porphyromonas endodontalis* are the top discriminators of COVID-19 disease severity (Figure 2D and E). Conversely, enrichment of *Bacteroides fragilis, Bacteroides caccae*, and *Clostridium clostridioforme* in the stool or another Bacteriodetes species: *Muribaculum intestinale* in the oral cavity are characteristic of individuals with moderate disease. We confirmed that most of the bacteria selected by the machine learning modeling were also different between the two groups by running differential expression analysis for sequence count data with DeSeq2 ^17^ (Supplementary Figure 1).

### SARS-CoV-2 antibody levels and patient outcomes

Patients with severe COVID-19 disease have been shown to have a different antibody trajectory compared to those with mild/moderate disease during hospitalization ^18^. Although, we only collected one blood sample per patient (at ∼ 6.42 ± 6.47 days after hospital admission) we aimed to investigate the relationship between patient microbiota and whether plasma levels of antibodies against the receptor binding domain (RBD) of the SARS-CoV-2 spike protein, specifically: IgA, IgM, and IgG varied depending on disease severity at time of blood collection or among patients who eventually succumbed to the disease vs. those who survived. We did not find any differences in antibody levels by disease severity (Figure 3A); however consistent with previous reports ^18^, we observed a significantly lower level of IgG against RBD in patients who eventually died (Figure 3B). High anti-RBD IgG antibody levels measured by ELISA have been shown to correlate with antibodies that block viral entry into host cells as measured neutralizing antibody assays ^19^.

**Figure 3.**
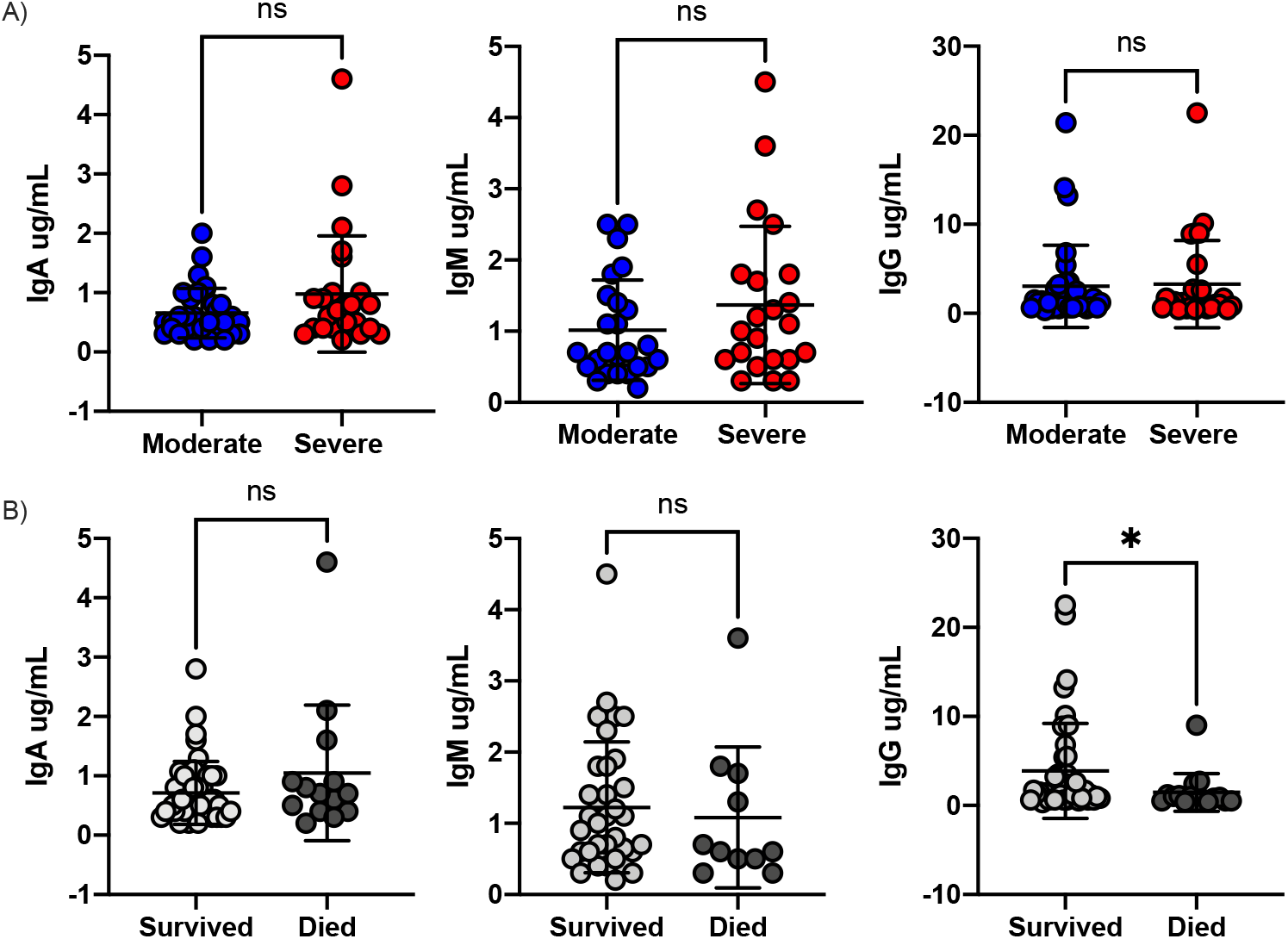
Antibody titers in COVID patients recruited in this study. A total of 54 blood samples were collected from patients that were hospitalized from April to June 2020. A) Antibody titers from patients with moderate (blue) and severe (red) COVID-19 symptoms. B) Antibody titers from patients that survived (light gray) or died (dark grey) to SARS-CoV-2 infection and. *p-value <0.05; Mann-Whitney test.

We further investigated associations between bacterial abundance and either IgA, IgM nor IgG antibody titers (Random Forest classification). We found no bacteria that predicted antibody levels, suggesting these are independent indicators of disease outcome.

## DISCUSSION

In this study, we have demonstrated that COVID-19 disease severity can be predicted by the stool or oral microbiome composition with higher accuracy than traditional clinical scoring methods using a combination of comorbidities and clinical biomarkers alone. Particularly, two pathobionts in the either the oral (*Porphyromonas endodontalis)* or intestinal (*Enterococcus faecalis*) microbiota can serve as indicator species to robustly predict the severity of SARS-CoV-2 infections. Our findings are clinically actionable as assessment of *Enterococcus faecalis* in feces will improve risk stratification of patients, which is highly relevant until this global pandemic is over. *Enterococcus faecalis* can be cultured from feces representing a cost-effective, rapid, and relatively easy test to implement in clinical settings. As such we urge the medical community that in addition to observable clinical variables, indicator species of the microbiome, specifically *Enterococcus faecalis*, can serve as robust predictor of COVID-19 severity and to quickly identify patients who are likely to require more supportive care or therapeutic interventions to improve chances of survival.

A hallmark of severe COVID-19 disease is an uncontrolled inflammatory response; specifically, a fulminant and fatal hypercytokinemia that causes an uncontrolled flood of immune cells into the lung ^20-22^. In these patients, it’s their own uncontrolled inflammatory response, rather than the virus, that causes severe lung injury and multi-organ failures leading to death. Gut microbial dysbiosis has been linked to activation of inflammatory immune networks that perpetuates several chronic diseases ^23,24^ (e.g., type 2 diabetes, hyperlipidemia), which are comorbidities associated with COVID-19 disease. Thus, a still unanswered question is the contribution of the microbiome in the immune response against SASR-CoV-2 infection.

A recent study has shown that SARS-CoV-2 infection triggers aberrant phenotypes of FoxP3+ T regulatory cells (Treg) ^25^, critical to immune homeostasis (reviewed ^26,27^). We and others have demonstrated the microbiota-dependent development and activation of Treg and their role in controlling exacerbated inflammatory responses ^4,28-36^. Thus, further studies with longitudinal sampling combined with analysis of Treg markers are needed to better understand how the dysbiosis in SARS-CoV-2 infected patients, and specifically the enrichment of the pathobionts we observed in this cohort, can contribute to COVID-19 disease severity via alteration of the Treg development.

## Data Availability

All sequencing data will be available upon submission.

## ACKNOWLEDGMENTS

All faculty members of the Microbiology and Physiological Systems Department and the EH& S at UMASS for insightful advice on sample containment. We are grateful for all the patients that participated in the study. We thank Katherine Fitzgerald for providing access to the BSL2+ laboratories to safely process all the samples. The Center for Microbiome Center, AM, GF, CF received funding to execute this study by the COVID-19/Pandemic Research Fund at UMass Medical School. Funding sources for AM, GF, CF were also provided by MassCPR Evergrande Award.

## METHODS

### Participant recruitment

We enrolled SARS-CoV-2 PCR positive patients with moderate (requiring < 4L of oxygen) or severe (requiring > 4L of oxygen) symptoms hospitalized at the University of Massachusetts Medical Center and UMASS Memorial Hospital from April 27 to June 10, 2020. This cohort was recruited under the COVID-COPE IRB protocol (docket # H00020145). The Institutional Review Board at the University of Massachusetts Medical School approved this study. Informed consent was obtained from all study participants or their health care proxy using RedCap digital signatures to reduce the potential for patient-staff transmission.

### Sample collection

All samples were collected by the doctor or nurse caring for the patient during standard of care rounds using all the necessary precautions. Stool samples were collected with a scoop from a paper stool catcher or directly from the ostomy bags into a sterile tube (Cat # 58-EZSAMPLER, ALPCO, NH, USA). Oral swabs were obtained using the OMNIgene•ORAL (DNAGenotek(tm), Canada) following the manufacturer instructions. Briefly, the tongue was swabbed for 30 seconds and then the swab was inserted into a tube with a DNA/RNA stabilizer buffer. For antibody assays, blood was collected in EDTA tubes (5 mL. BD Vacutainer® tubes, Becton Dickinson, USA) using sterile technique and blood borne pathogen precautions enhanced for COVID-19 patients. Plasma was separated from peripheral blood cell pellet by centrifugation, 10 minutes, room temperature and aliquots stored at −20 °C until thawed for ELISA testing.

### Clinical data

All the clinical data was obtained retrospectively by reviewing medical records of each participant.

### DNA and RNA isolation

Prior to isolation, SARS-CoV-2 was inactivated in all samples by heat at 65-70 °C for one hour as done elsewhere ^37,38^. After viral deactivation, nucleic acid isolation for stool and oral samples was performed using the ZymoBIOMICS DNA/RNA Miniprep Kit (Cat # D7003/D7003T, Zymo Research, CA, USA) following the manufacturer recommendations for parallel isolation of DNA and RNA. Oral samples were first treated with the addition of 5ul Proteinase K (Cat # P8107S, New England Biolabs, MA, USA) and incubated for 2 hours at 50 °C. 250ul of the treated sample was used for extraction. Extraction of total RNA from blood samples was performed using the Tempus(tm) Spin RNA isolation kit (Cat # 4380204, Applied Biosystems, USA) following manufactures instructions.

### Microbiome profiling

The *16S rRNA* gene was sequenced following methods previously described ^39^ using the 341F and 806R universal primers to amplify the V3-V4 region. The 300nt paired-end sequences were generated on the Illumina MiSeq platform. Replicate reactions were performed for each sample and the read data was combined in analysis. Forward and reverse 16S MiSeq-generated amplicon sequencing reads were dereplicated and sequences were inferred using dada2 ^40^. Potentially chimeric sequences were removed using consensus-based methods. Taxonomic assignments were made using BLASTN against the NCBI refseq rna database. These files were imported into R and merged with a metadata file into a single Phyloseq object.

### Mathematical modeling

#### Machine learning analysis to predict outcome from microbiome and clinical covariates

We run random forest classification (RFC) to identify stool, oral bacteria and clinical covariates that are predictive of clinical outcome SARS-CoV-2 disease fatality and severity. Our RFC pipeline consists in a first step of feature selection in where the wrapper Boruta ^41^ is used to determine a subset of covariates that is predictive of the outcome, and a second step in where RFC is run using only the Boruta-selected subset. To estimate the accuracy in predicting clinical outcome we used leave-one-out cross-validation scheme and corresponding Area Under the Curve (AUC). AUC values were used to compare different models in terms of prediction accuracy. To interpret the results from the RFC analysis, the RFC models were input into Local Interpretable Model-agnostic Explanation (LIME) toolbox ^42^. LIME trains a local surrogate model that explains the predictions of black-box machine learning model such as Random Forest. In our context LIME identifies human-interpretable logical rules on the microbiome that discriminate between patients with different outcomes (e.g., *abundance of bacterium X less that normalized count K is characteristic of Severity Moderate*). The LIME output was used to determine the prevalence of a rule (e.g., in how many cross-validations a rule was selected) and the number of times it contributes to predicting the correct label. This computational scheme was used to predict SARS-CoV-2 disease fatality and severity (binary variables). The clinical covariates corresponding to categorical variables were one-hot-encoded. For microbial abundances we used the Amplicon Sequence Variant (ASV) counts normalized using DeSeq2 ^43^.

#### Differential analysis to confirm microbiota-outcome associations

To confirm the associations between stool and oral bacteria and clinical outcome we run Differential expression analysis for Sequence count data in DeSeq2 as done previously ^44^. Specifically, we run the model *Counts ∼ Phenotype + CC* (where *CC* stands for the main clinical covariates selected by the only CC model). ASVs with Benjamin-Hochberg adjusted p value less than 0.05 for the Phenotype variable were considered differential.

### Antibody ELISA

Antibodies against the receptor binding domain (RBD) of the SARS-CoV-2 spike protein were measured by ELISA following published methods ^45^. In brief, IgG, IgA and IgM antibodies against the RBD recombinant protein (gifted from MassBiologics) was used at 0.5 µg/mL and incubated with plasma at a 1:100 and 1:1000 dilution. Absorbance was measured at 450 nm and 570 nm on the SpectraMax iD5 ELISA plate reader (Molecular Devices) using SoftMax Pro software (version 7.1, Molecular Devices). For the positive antibody control, CR3022 was diluted from a concentration of 2.5 μg/ml in dilution buffer to 12 two-fold serial dilutions to generate the standard control curve. The 570 nm OD was subtracted from the 450 nm OD for the final OD value. Antibody levels were used as a continuous variable in the analysis.

### Statistical analysis

Fisher T-test, Chi-square, and Mann-Whitney tests were used to evaluate differences in demographics and antibiotic use among patients with moderate *vs*. severe COVID-19 disease. A non-parametric Mann-Whitney U-test was also used to evaluate differences in antibody titer in serum by disease severity or clinical outcome.

---

**Supplementary Table 1.** Clinical variables included in the models obtained by review of medical records.

| **Clinical variables included in the models** | **Moderate**<br>**(n=32)** | **Severe**<br>**(n=31)** | **Mann Whitney**<br>**tests (p value)** |
| --- | --- | --- | --- |
| *Symptoms* |  |  |  |
| Abdominal pain | 1 | 1 | >0.999999 |
| Chest pain | 2 | 3 | 0.671867 |
| Chills | 5 | 6 | 0.74996 |
| Cough | 12 | 12 | >0.999999 |
| Diarrhea | 3 | 4 | 0.707846 |
| Difficulty breathing | 16 | 22 | 0.12349 |
| Dizziness | 1 | 0 | >0.999999 |
| Dyspnea | 14 | 19 | 0.210129 |
| Fever | 11 | 17 | 0.131294 |
| Hemoptysis | 0 | 1 | 0.492063 |
| Hypotension | 4 | 2 | 0.671867 |
| Hypoxic respiratory distress | 18 | 25 | 0.057677 |
| Shortness of breath | 16 | 21 | 0.202826 |
| Sore throat | 1 | 0 | >0.999999 |
| Tachycardia | 9 | 11 | 0.595035 |
| Tachypnea | 11 | 17 | 0.131294 |
| Vomiting | 4 | 2 | 0.671867 |
| Anemia | 3 | 5 | 0.474123 |
| *Co-morbidities* |  |  |  |
| Asthma | 7 | 2 | 0.147716 |
| Bradycardia | 2 | 0 | 0.492063 |
| Cardiomyopathy | 3 | 2 | >0.999999 |
| Cancer | 6 | 8 | 0.556127 |
| Cerebellar atrophy | 0 | 0 | >0.999999 |
| Cerebral palsy | 1 | 0 | >0.999999 |
| Chronic fibrillation | 6 | 8 | 0.556127 |
| Chronic anticoagulation | 3 | 6 | 0.302018 |
| Heart failure | 8 | 12 | 0.286909 |
| Kidney disease | 7 | 8 | 0.773534 |
| Chronic respiratory failure | 2 | 7 | 0.081587 |
| Thrombosis | 4 | 1 | 0.354671 |
| Cirrhosis | 1 | 1 | >0.999999 |
| Congestive heart failure | 7 | 12 | 0.176864 |
| COPD | 3 | 7 | 0.183638 |
| Coronary artery disease | 2 | 8 | 0.043318 |
| Emphysema | 0 | 2 | 0.238095 |
| End stage renal disease | 2 | 5 | 0.248622 |
| Epilepsy | 2 | 4 | 0.425766 |
| Goodpasture syndrome | 1 | 0 | >0.999999 |
| Heart attack | 0 | 1 | 0.492063 |
| Heart failure | 8 | 13 | 0.187691 |
| Hepatic encephalopathy | 1 | 0 | >0.999999 |
| History (hx) adenocarcinoma | 1 | 3 | 0.354671 |
| History (hx) aortic aneurysm | 1 | 0 | >0.999999 |
| History (hx) cancer | 6 | 8 | 0.556127 |
| Hypercholesteremia | 1 | 11 | 0.001153 |
| Hyperglycemia | 13 | 15 | 0.61593 |
| Hyperlipidemia | 16 | 13 | 0.620886 |
| Hypertension | 23 | 26 | 0.36492 |
| Hypotension | 0 | 1 | 0.492063 |
| Interstitial lung disease | 0 | 1 | 0.492063 |
| Lymphoma | 1 | 0 | >0.999999 |
| Multiple myeloma | 1 | 1 | >0.999999 |
| Nephrectomy | 0 | 1 | 0.492063 |
| Parkinson's | 0 | 1 | 0.492063 |
| Renal disease | 7 | 8 | 0.773534 |
| Renal failure | 2 | 5 | 0.256519 |
| Stage 3 chronic kidney disease | 4 | 4 | >0.999999 |
| Stage 4 adenocarcinoma | 1 | 1 | >0.999999 |
| Stage 5 end stage renal disease | 2 | 5 | 0.256519 |
| Type 1 diabetes | 0 | 1 | 0.492063 |
| Type 2 diabetes | 13 | 15 | 0.61593 |

**Supplementary Table 2.** Performance metrics of different Random Forest Classification Models predicting COVID-19 severity (moderate *vs*. severe). SEN=Sensitivity, SPEC=Specificity, PPV=Positive Predicted Values, NPV=Negative Predicted Values, PREC=Precision, REC=Recall, F1=F1 Score.

| ***MODEL*** | **SEN** | **SPEC** | **PPV** | **NPV** | **PREC** | **REC** | **F1** |
| --- | --- | --- | --- | --- | --- | --- | --- |
| ***CC*** | 0.68 | 0.68 | 0.68 | 0.68 | 0.68 | 0.68 | 0.68 |
| ***STL*** | 0.86 | 0.85 | 0.86 | 0.85 | 0.86 | 0.86 | 0.86 |
| ***TNG*** | 0.72 | 0.87 | 0.84 | 0.76 | 0.84 | 0.72 | 0.78 |
| ***CC + STL*** | 1.00 | 0.80 | 0.85 | 1.00 | 0.85 | 1.00 | 0.92 |
| ***CC + TNG*** | 0.76 | 0.90 | 0.88 | 0.79 | 0.88 | 0.76 | 0.81 |

**Supplementary Figure 1:**
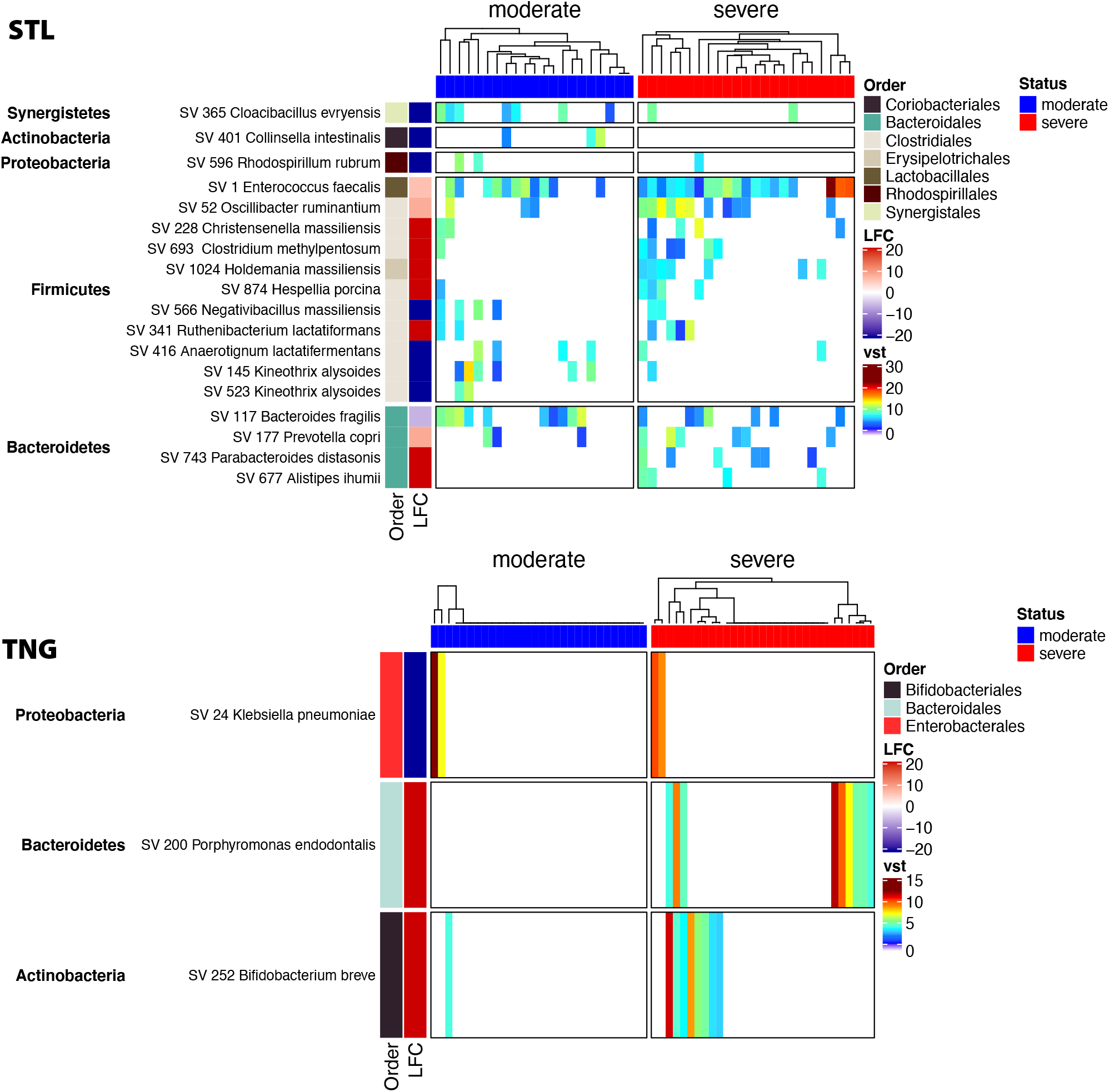
Differential Analysis results from DeSeq2. DeSeq2 was used to fit the model *Counts ∼ Severity* for the stool (STL) or oral (TNG) microbiomes. Heatmap display Amplicon Sequence Variants (ASVs) with significant *p value* (BH corrected *p value* < 0.05).

